# Long-Term Clinical Outcomes in Critically Ill Patients with Sepsis and Pre-existing Sarcopenia: A Retrospective Cohort Study

**DOI:** 10.1101/2023.04.12.23288490

**Authors:** Nola Darden, Sonakshi Sharma, Xue Wu, Benjamin Mancini, Kunal Karamchandani, Anthony S Bonavia

## Abstract

**Purpose:** Critically ill patients with sepsis account for significant disease morbidity and healthcare costs. Sarcopenia has been proposed as an independent risk factor for poor short-term outcomes, although its effect on long-term outcomes remains unclear.

**Methods:** Retrospective cohort analysis of patients treated at a tertiary care medical center over 6 years (09/2014 - 12/2020). Critically ill patients meeting Sepsis-3 criteria were included, with sarcopenia defined by skeletal muscle index at the L3 lumbar area on abdominal Computed-Tomography scan. The prevalence of sarcopenia and its association with clinical outcomes was analyzed.

**Results:** Sarcopenia was present in 34 (23%) of 150 patients, with median skeletal muscle indices of 28.1 cm^2^/m^2^ and 37.3 cm^2^/m^2^ in sarcopenic females and males, respectively. In-hospital mortality was not associated with sarcopenia when adjusted for age and illness severity. One year mortality was increased in sarcopenic patients, after adjustment for illness severity (HR 1.9, p = 0.02) and age (HR 2.4, p = 0.001). However, it was not associated with increased likelihood for discharge to long-term rehabilitation or hospice care in adjusted analyses.

**Conclusion:** Sarcopenia independently predicts one year mortality but is not associated with unfavorable hospital discharge disposition in critically ill patients with sepsis.

## INTRODUCTION

Since primary sarcopenia (decreased ‘reserve of strength’) results from normal aging, it often coexists with other age-related diseases such as cancer and sepsis [1-3]. Sepsis, defined as life-threatening organ dysfunction caused by a dysregulated host response to infection [4], remains the leading cause of death in hospitalized patients [5-7]. Particularly adverse clinical outcomes are observed in septic patients who are older than 65 years [8]. Advances in critical care therapy, however, have caused an increasing proportion of aged, sepsis survivors to live longer and experience significant, previously unseen morbidity including long-term rehabilitation and decreased quality of life [9-11]. The frequent coexistence of sarcopenia and sepsis has, thus, created a need to better understand how these conditions interact to influence patient outcomes. This knowledge may allow clinicians to plan for appropriate rehabilitation and nursing care, early in course of the hospitalization in high-risk patients. It may also facilitate discussions by clinicians with patients and their families regarding anticipated disease prognosis and long-term goals of medical care.

For practical and research purposes, sarcopenia is often defined in terms of skeletal muscle index, that is, skeletal muscle area (measured on Computed-Tomography (CT) imaging) indexed to patient height [12]. While the pathophysiology of primary sarcopenia is not completely understood [13], it involves changes in both muscle size and muscle quality. Specifically, sarcopenia is believed to be caused by a reduced number of myofibers, hypertrophy of remaining fibers, infiltration of muscle with adipose tissue and a decreased numbers of satellite cells, which are responsible for adult myogenesis [13-15]. In later stages of the disease, decreased muscle generation is coupled with fibrosis, resulting in profound structural and functional muscular impairment.

The first study that investigated the role of sarcopenia in septic patients was conducted in 2017, reporting that decreased skeletal muscle mass was associated with higher in-hospital mortality in older patients with sepsis [16]. A more recent study, using similar methods and cohort size, concluded that muscle wasting associated co-morbidities, rather than sarcopenia itself, were risk factors for in-hospital mortality [17]. The limitation of both studies is the short interval to patient follow-up. Both measured in-hospital mortality as their primary outcome, with Baggerman *et al* reporting a median (IQR) hospital length of stay of 21 (11-38) days [17]. In contrast, current literature surrounding chronic, post-sepsis illness suggests that much of the disease morbidity occurs in the weeks to months following acute illness [18]. A meta-analysis of 2396 patients investigated sarcopenia as a risk factor for poor post-sepsis outcomes, similarly reported early (in-hospital or 1-month) mortality as the primary measured outcome [19]. This meta-analysis incorporated studies utilizing three different definitions of sepsis, the earliest of which dates back to 1992 [20].

The importance of investigating sarcopenia as an independent risk factor for poor post-septic outcomes is underlined by aging global populations and the resultant demographic of patients who are increasingly seeking healthcare [21]. In the present study, we hypothesized that sarcopenia would adversely predict *long-term* outcomes in critically ill patients with sepsis. To investigate our hypothesis, we performed a retrospective analysis of these patients, as defined by the Sepsis-3 criteria [4]. Besides one-year mortality as the primary outcome, we investigated the effect of sarcopenia on ‘poor’ post-hospitalization discharge disposition. We used the latter as a surrogate marker of quality-of-life following patients’ index hospitalization. ‘Poor’ discharge disposition was defined as transfer to a long-term rehabilitation or nursing facility, rather than home.

## METHODS

### Study Design

This was a single-center, retrospective cohort analysis of patients hospitalized at a tertiary care medical center between 09/2014 and 12/2020. Adults (>18 years old) who were critically ill and had acute-onset sepsis were included. Given the known limitations of using administrative data such as *International Classification of Diseases, Ninth Edition, Clinical Modification* (ICD-9-CM) sepsis codes for identifying actual incidences of this disease [22, 23], a research investigator manually ascertained which of the identified patients met study inclusion criteria. A diagnosis of sepsis required that a patient experienced a change in sequential organ failure assessment (SOFA) score of two or more, in the setting of clinically suspected or microbiologically proven infection, in accordance with the Sepsis-3 Criteria [4]. Critical illness was defined by billing data consisting of Centers for Medicare and Medicaid Services (CMS), Current Procedural Terminology (CPT) codes of 99291 or 99292. Patients who did not have a Computed-Tomography (CT) scan of their abdomen performed within 7 days of index hospitalization were excluded. The manuscript was written according to the Strengthening the Reporting of Observational studies in Epidemiology (STROBE) criteria [24].

### Assessment of Sarcopenia

Axial CT scan images at the L3 vertebral level were obtained from the Picture Archiving and Communication System (PACS) in Digital Imaging and Communication in Medicine (DICOM) format for all patients. Sarcopenia was defined as ≤5^th^ percentile of skeletal muscle area index of a gender-matched, reference population (41.6 cm^2^/m^2^ for males and 32.0 cm^2^/m^2^ for females), as previously described [17, 25]. Abdominal and/or pelvic CT imaging performed within 7 days of the date of index hospitalization were used to define sarcopenia. Specifically, skeletal muscle area was measured on axial imaging of the L3 spinal level, using ImageJ software (version 1.53t, U. S. National Institutes of Health, Bethesda, Maryland, USA)[26], as illustrated in **Fig 1** and using methods previously described [27]. Both CT images obtained with and without intravenous contrast were used in the calculation of skeletal muscle area, as previously reported [28, 29].

**Figure 1.**
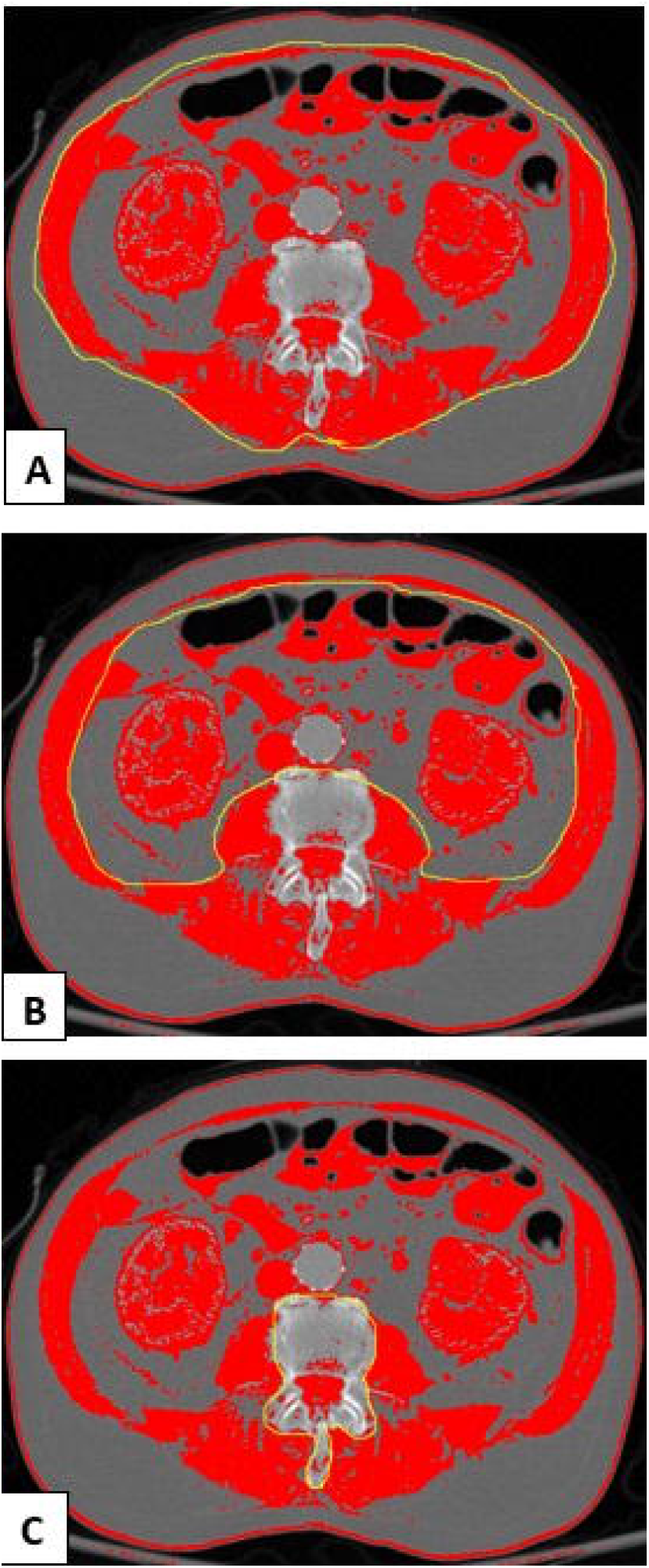
Measuring skeletal muscle on L3 axial slice Computer-Tomography (CT) Image, Using the National Institutes of Health ImageJ. Radio density thresholds were selected to correlate with those of skeletal muscle, with lower and upper thresholds of −29 and 150 Hounsfield units, respectively. (A) Outer perimeter of the abdominal muscles is first delineated (yellow tracing) and area calculated, (B) Process is repeated for the inner perimeter of abdominal muscles (yellow tracing), (C)Vertebral area is further excluded (yellow tracing). The difference between measures equates to the area of skeletal muscle at the L3 level.

### Clinical Outcomes

Demographic data, medical comorbidities and short- and long-term clinical outcomes were obtained from the electronic medical record. Medical comorbidities were identified using ICD-9 billing codes. Severity of acute illness was defined by Acute Physiology and Chronic Health Evaluation (APACHE) II and Sequential Organ Failure Assessment (SOFA) scores, which were calculated by the research investigator [30, 31].

### Statistical Analysis

Our sample size of 150 patients was based on two recent, comparable, retrospective cohort studies that assessed the effects of sarcopenia on short-term, clinical outcomes [16, 17]. For binary and nominal outcomes, we constructed frequencies and percentages as descriptive statistics, and implemented Fisher’s exact test statistics to compare patients with and without sarcopenia. For continuous outcomes, we constructed median and inter-quartile ranges (IQR) as descriptive statistics and implemented Wilcoxon rank-sum tests to compare groups with and without sarcopenia. One-year survival was defined as the time interval between admission/discharge date and last visit or death within 365 days, which we analyzed via Kaplan-Meier survival curves and log-rank tests. Survival data was truncated at one year follow-up to ensure equivalent comparison of long-term outcomes between patients. We performed two subsequent analyses of our outcome data to account for potential confounding variables. We applied a Cox proportional hazards regression analysis to assess the effect of sarcopenia on one-year survival, corrected for variables known to be related to illness severity, such as SOFA and APACHE II. In addition, we applied a multivariable logistic regression analysis to assess the effect of sarcopenia on discharge disposition, adjusted for illness severity. Analyses were performed using R v4.1.2 (R core team, 2022).

## RESULTS

### Study Population

Of the 150 patients that were included, 34 patients (23%) met criteria for pre-existing sarcopenia within the 6-year study period (**Fig 2**). Patients with sarcopenia were older and had a lower body mass index as compared with non-sarcopenic patients (**Table 1**). Median SOFA and APACHE II scores for patients with sarcopenia were higher than those without sarcopenia (p = 0.025 and 0.019, respectively).

**Table 1.**
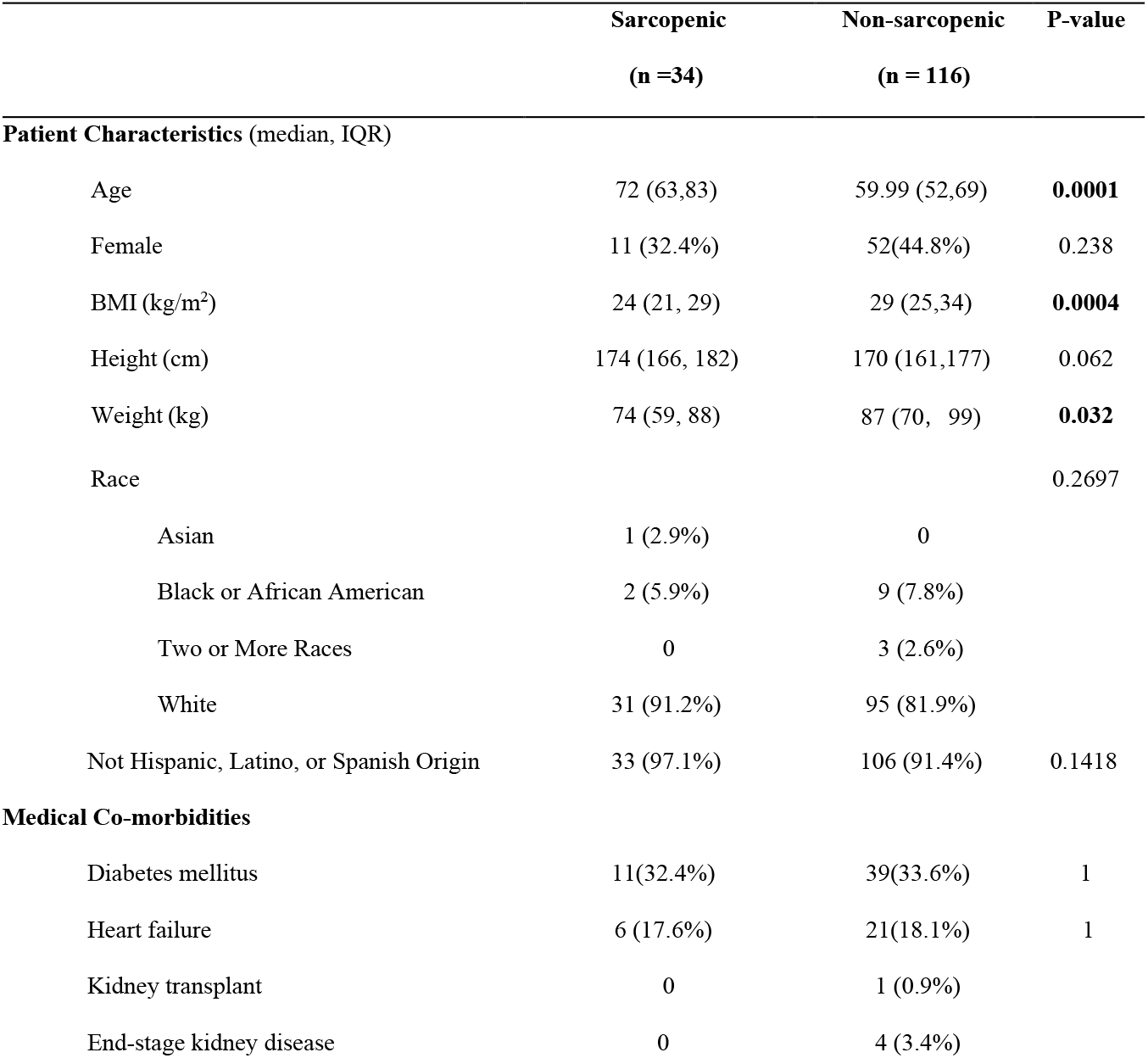
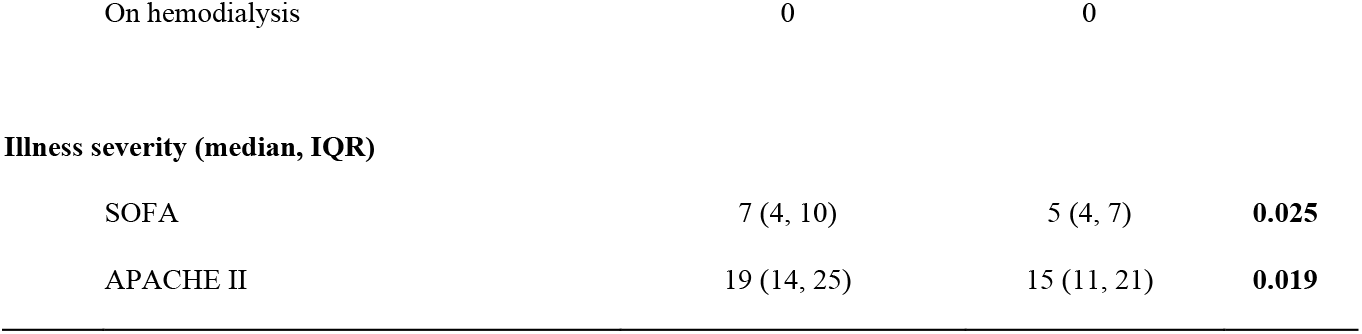
Demographic and Clinical Profile

**Figure 2.**
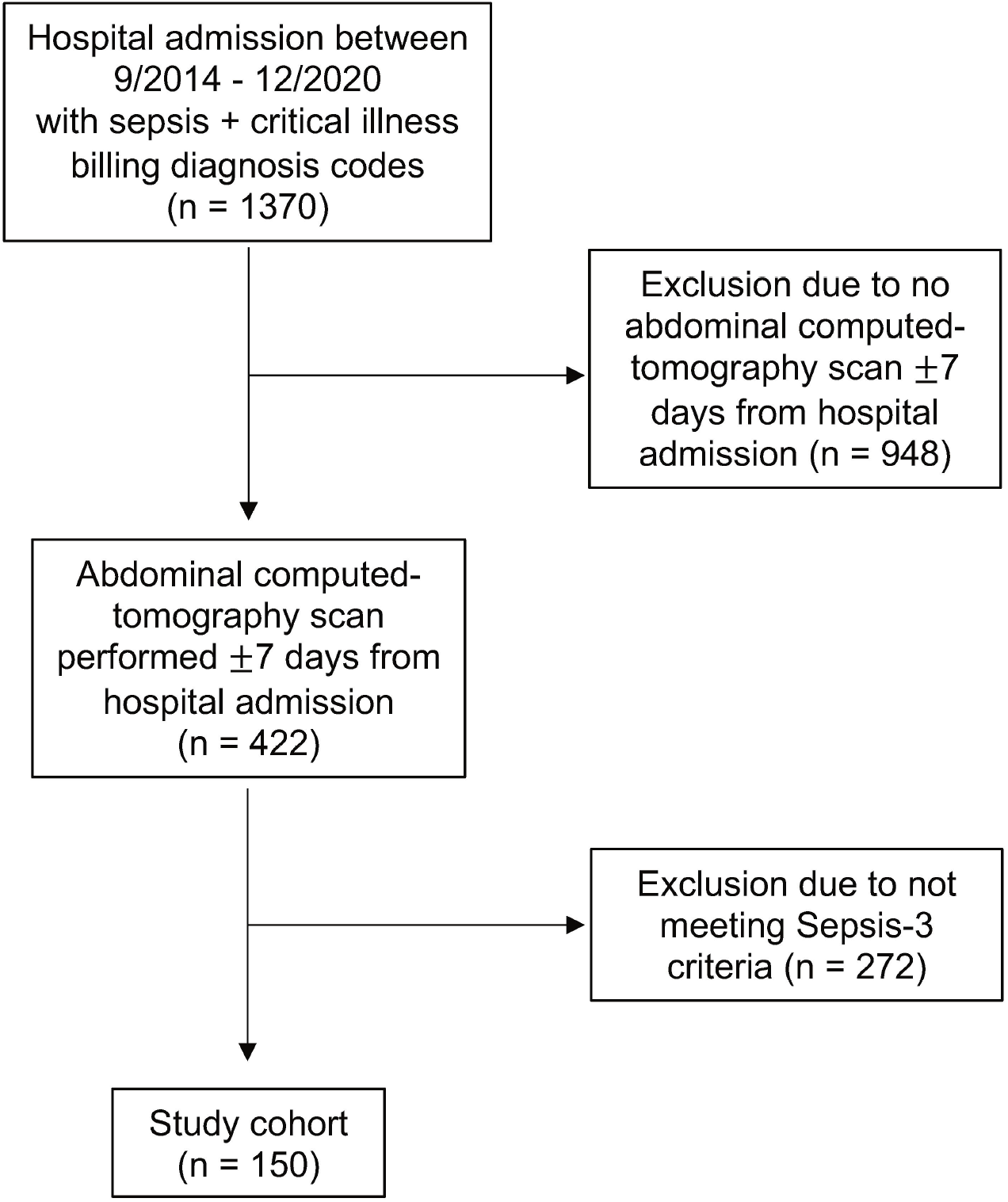
Flow diagram demonstrating numbers of individuals included and excluded at each stage of the study.

### Distribution of Sarcopenia

Sex- and age-specific body composition parameters are shown **Table 2**. Amongst sarcopenic patients, 11 (32.4%) were female and 31 (91.2%) were white. Median (IQR) skeletal muscle area for females was 73.7 cm^2^ (69.3 – 81.5 cm^2^) and skeletal muscle index was 28.1 cm^2^/m^2^ (26.1 – 30.3 cm^2^/m^2^). The median skeletal muscle area for sarcopenic males was 119.0 cm^2^ (104.8 – 123.9 cm^2^) and skeletal muscle index was 37.3 cm^2^/m^2^ (33.7 – 40.0 cm^2^/m^2^). Amongst non-sarcopenic patients, median (IQR) skeletal muscle area for females was 113.7 cm^2^ (100.9 – 127.8 cm^2^) and skeletal muscle index was 44.8 cm^2^/m^2^ (39.8 – 49.1 cm^2^/m^2^). The median skeletal muscle area for sarcopenic males was 155.6 cm^2^ (143.5 – 176.4 cm^2^) and skeletal muscle index was 49.1 cm^2^/m^2^ (45.5 – 57.6 cm^2^/m^2^).

**Table 2.**
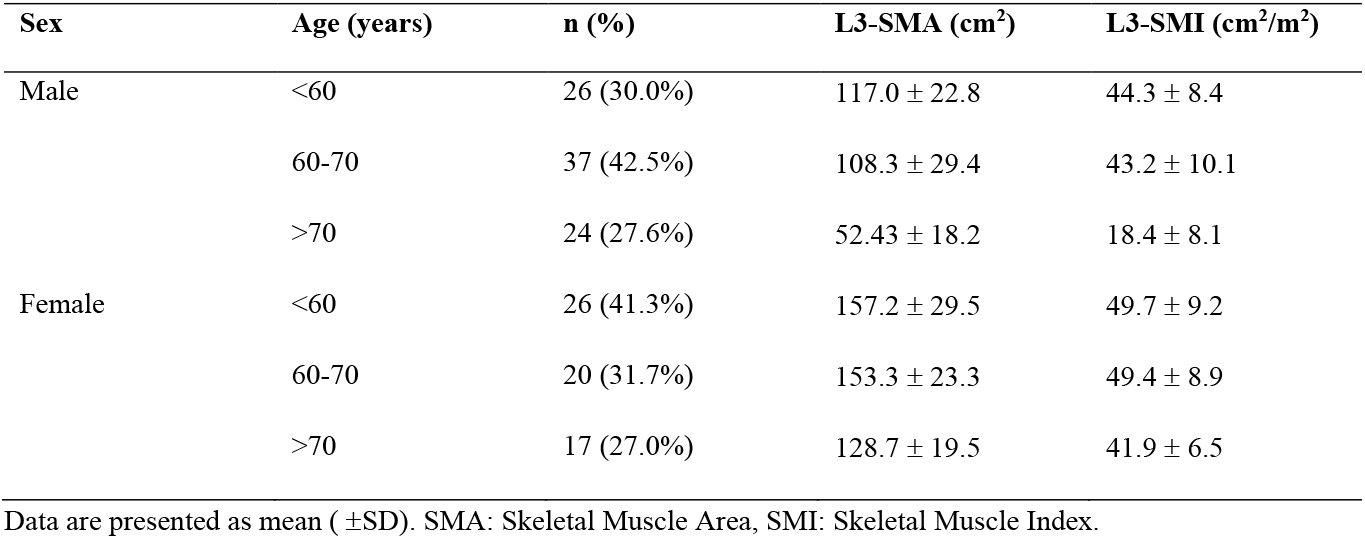
Body composition of critically ill patients with sepsis

### Association between sarcopenia and in-hospital mortality

Univariable analysis of clinical outcomes in sarcopenic and non-sarcopenic patients is shown in **Table 3**. Sarcopenia did not significantly affect the in-hospital mortality rate, with an odds ratio of 2.03 (95% CI 0.8 – 4.9, p = 0.121) (**Fig 3**). Given that SOFA and APACHE II scores have been shown to predict mortality in critically ill patients [30, 32, 33] and differed between sarcopenic and non-sarcopenic cohorts (**Table 1**), we performed two subsequent logistic analyses to account for these confounding variables. Consistent with previous studies, when illness severity was factored into the analysis, risk of in-hospital mortality with sarcopenia remained non-significant (**Fig 4A, B**) [17].

**Table 3.**
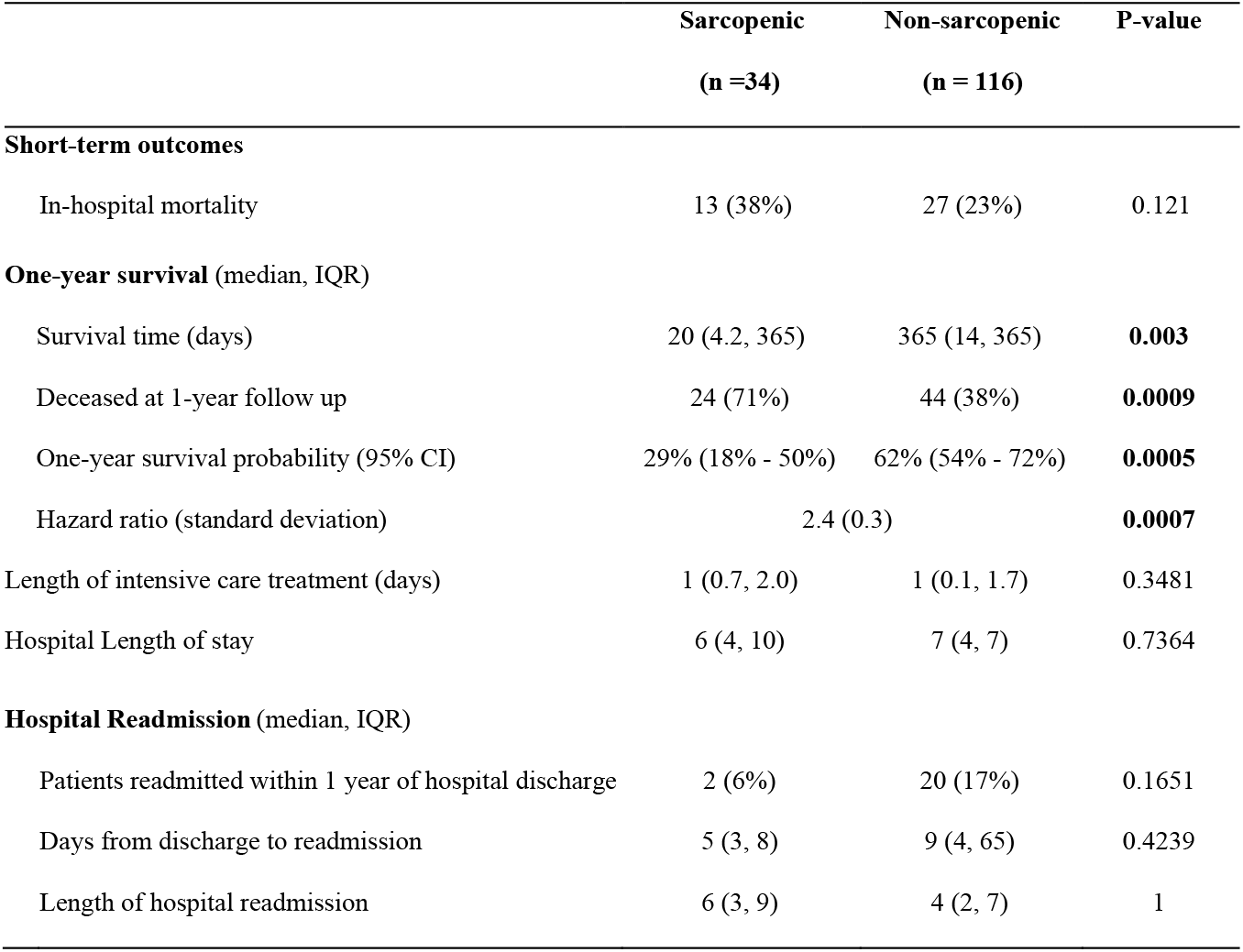
Clinical outcomes in sarcopenic and non-sarcopenic patients

**Figure 3.**
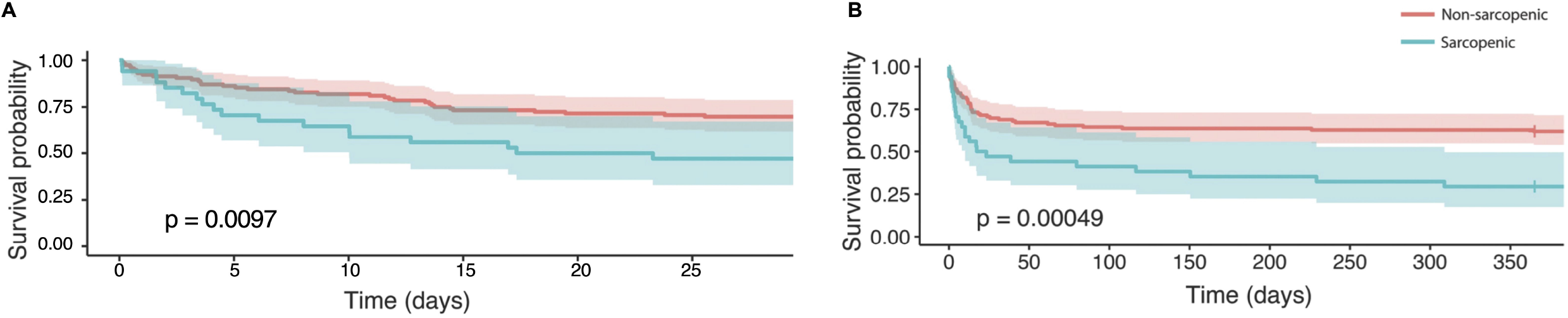
Kaplan-Meier survival curves demonstrating the effect of sarcopenia on (A) in-hospital mortality, and (B) 1 year mortality, in septic patients. 95% confidence intervals are shown.

**Figure 4.**
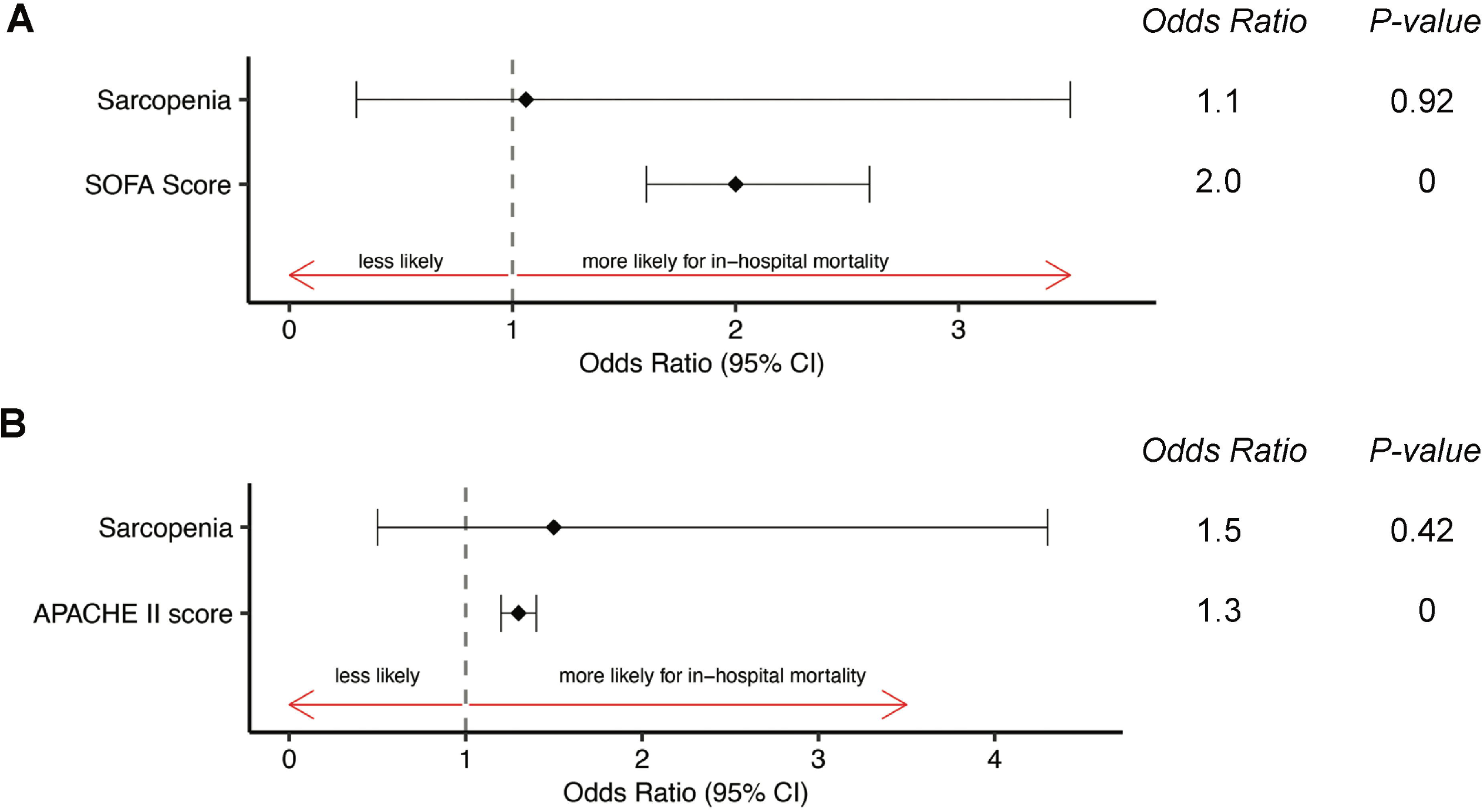
Logistic analysis, assessing the effect of Sarcopenia on in-hospital mortality when adjusting for severity of illness by (A) SOFA score and (B) quartiles of APACHE II score.

### Association between sarcopenia and one year survival

In univariable analysis, sarcopenia significantly decreased one-year survival, with an estimated survival probability of 29.4% in sarcopenic patients (hazard ratio 2.34; 95% CI: 17.5% - 49.5%; p = 0.0005) (**Table 3, Fig 3B**). When adjusting for SOFA score by Cox multivariable regression, sarcopenia remained an independent risk factor for one-year mortality (hazard ratio 1.8; 95% CI 1.1 – 3.0; p = 0.02) (**Fig 5A**). APACHE II score was utilized as an alternative mortality-risk indicator for severity of illness adjustment, with the lowest scoring quartile (APACHE II scores of 2 to 11) used as the reference group. Cox multivariable regression revealed that sarcopenia remained an independent risk factor for one-year survival despite adjustment for APACHE II score (hazard ratio 1.9, 95%CI: 1.1 – 3.1, p = 0.02) (**Fig 5B**).

**Figure 5.**
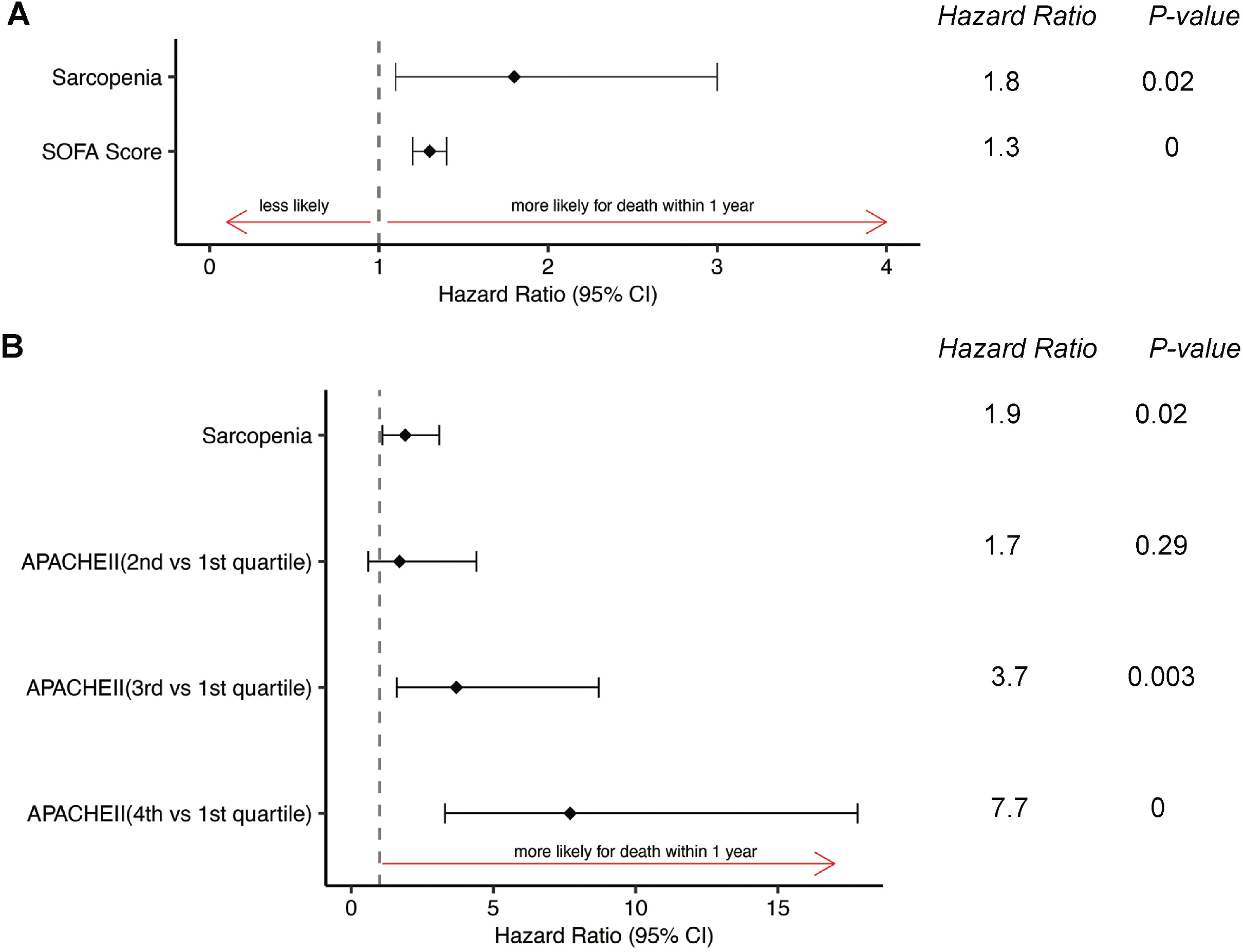
Cox regression analysis, assessing the effect of sarcopenia on one-year survival when adjusting for severity of illness by (A) SOFA score and (B) quartiles of APACHE II score.

### Age adjustment for risk of adverse outcomes in sarcopenic patients

Sarcopenia can either be primary (age-related) or secondary to chronic illnesses such as cardiorespiratory failure, diabetes mellitus, immobilization or chronic inflammatory disease [34]. Since the chronic illnesses that account for secondary sarcopenia are also more prevalent in older patients [35], we analyzed the age-adjusted effects of sarcopenia on outcomes. We used an age cutoff of 70 years to define ‘older’ versus ‘younger’ patients with sepsis, based on data from previous epidemiologic studies [8, 36, 37]. While the incidence of sarcopenia was significantly lower in younger, septic patients (odds ratio 0.263), univariable Cox analysis of one-year mortality in 48 older patients demonstrated that age itself is not a significant predictor of one-year survival following sepsis (**Table 4**). When adjusted for age, the hazard ratio associated with sarcopenia alone was 2.39 (p<0.05).

**Table 4.**
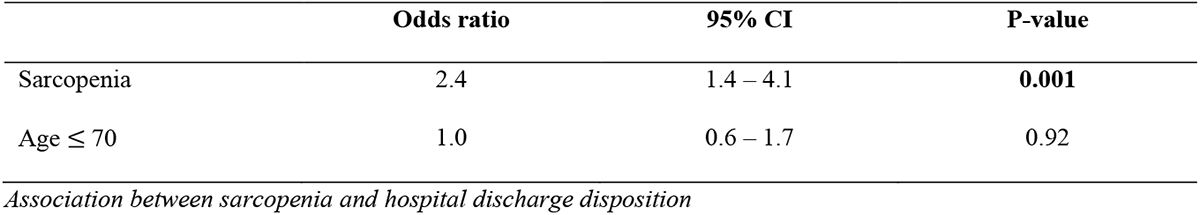
Univariable Cox analysis, assessing the effects of sarcopenia and age on one-year survival

### Association between sarcopenia and hospital discharge disposition

‘Favorable’ discharge disposition was defined as transfer of sepsis survivors from hospital directly to their home, while ‘unfavorable’ discharge disposition was defined as transfer from hospital to a rehabilitation facility (skilled nursing facility or inpatient rehabilitation facility) or hospice care. Univariable logistic regression analysis revealed that the proportion of patients experiencing unfavorable post-hospital discharge disposition was higher in the sarcopenic group, affecting 67% of these patients as compared with 37% of non-sarcopenic patients (odds ratio 3.4; 95% CI: 1.3 – 9.8; p = 0.02) (**Table 5**).

**Table 5.**
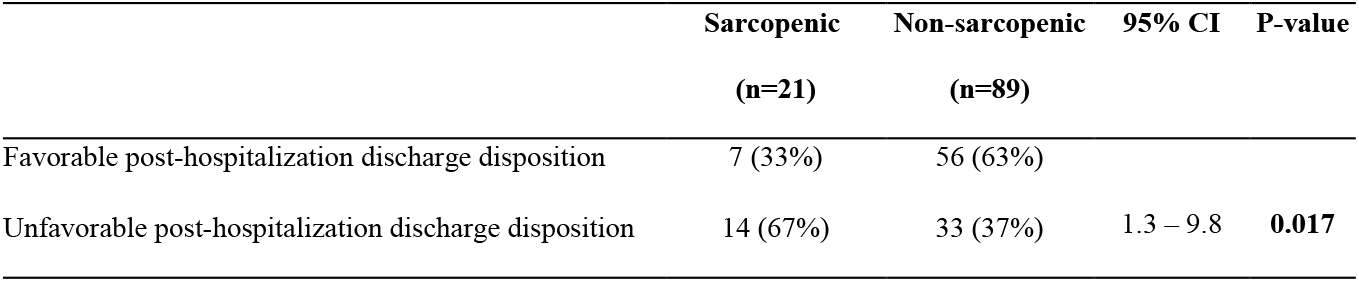
Univariable logistic analysis of sarcopenia and hospital discharge disposition

In multivariable logistic regression adjusted for SOFA score, the odds of unfavorable hospital discharge were 3.1 times higher in sarcopenic patients (p = 0.03) (**Fig 6A**). After adjustment for APACHE II score, the effect of sarcopenia on unfavorable hospital discharge disposition no longer remained significant (odds ratio: 2.9; 95% CI: 1.0 – 9.3; p = 0.06) (**Fig 6B**). Similarly, while univariable logistic analysis demonstrated that older, septic patients are more likely to be discharged to a long-term nursing facility or hospice care (odds ratio 0.225, p = 0.0005), age- adjusted analysis did not find a significant correlation between sarcopenia and unfavorable discharge disposition (odds ratio 2.31, p = 0.126).

**Figure 6.**
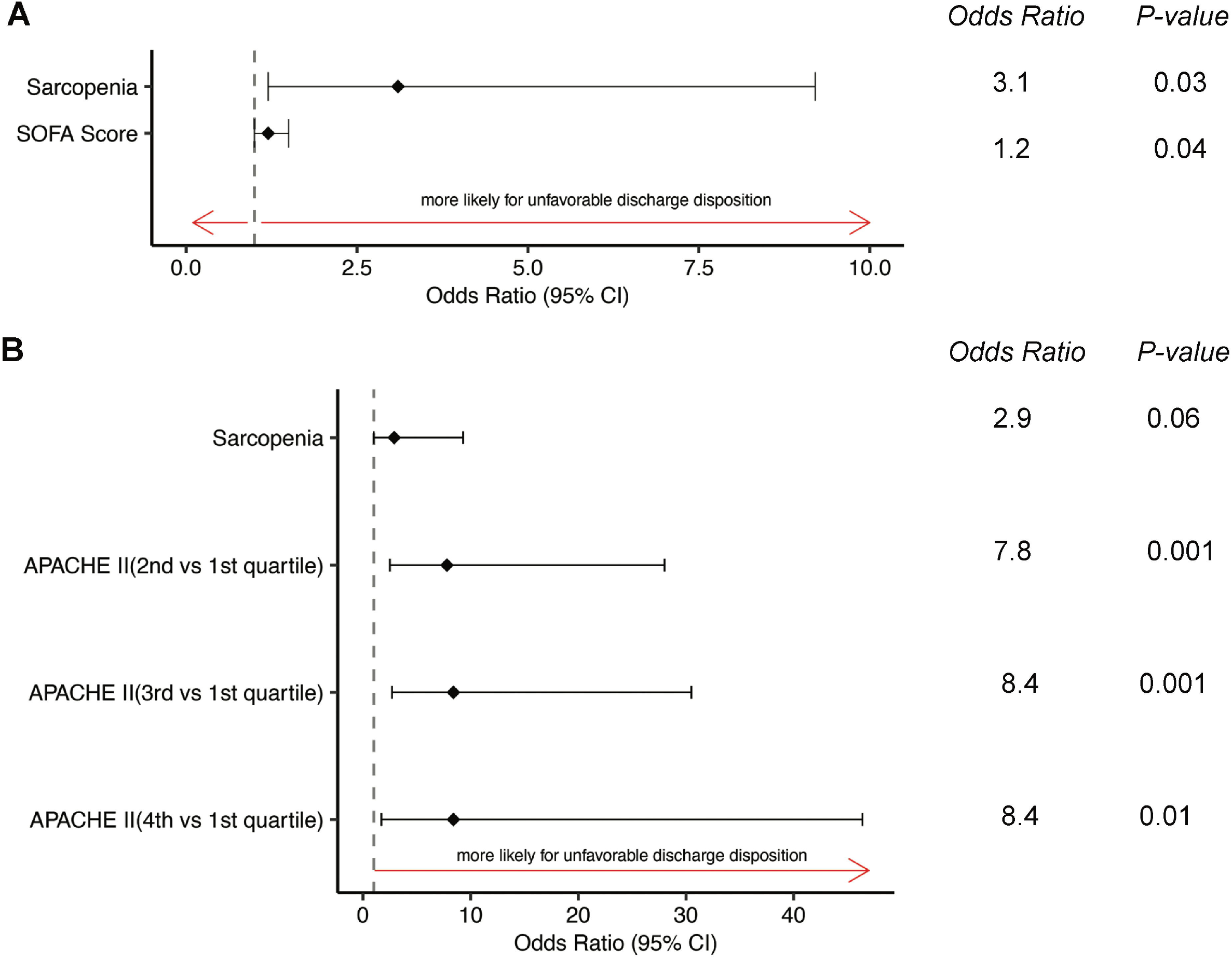
Logistic analysis, assessing the effect of assessing the effect of sarcopenia on one-year survival when adjusting for severity of illness by (A) SOFA score and (B) APACHE II score. Unfavorable discharge disposition was defined as transfer from hospital to a rehabilitation facility (skilled nursing facility or inpatient rehabilitation facility) or hospice care.

## DISCUSSION

Declining mortality rates in acute sepsis, resulting from faster disease recognition and implementation of sepsis bundles, have been hailed a success of modern medicine [38-40]. However, the huge healthcare costs of sepsis management ($62 billion annually, and highest amongst admissions for all disease states), combined with the large proportion of patients developing prolonged illness and indolent death within a year, make these decreases in short-term mortality rates appear less impressive [5, 6, 41, 42]. Rapid and accurate assessments of sarcopenia and frailty have gained increasing traction amongst clinicians seeking to risk-stratify patients during their hospitalization or prior to elective surgery [43, 44]. These measures of muscle dysfunction have also been proposed to explain the discordance between decreased inpatient mortality and persistently poor, one-year outcomes [45]. In this retrospective cohort analysis, we observed a high prevalence of pre-existing sarcopenia in critically ill patients with sepsis. We also identified sarcopenia as an independent risk factor for one-year mortality, although sarcopenia itself did not increase the risk for unfavorable hospital discharge disposition in age-adjusted analysis.

Low muscle mass has been shown to predict in-hospital and short-term mortality in critically ill patients, although its effects on longer-term disease outcomes have not been extensively explored [46]. This distinction between short- and long-term outcomes merits emphasis, since it highlights the paradigm shift in sepsis from a disease that is often deadly in the short-term to one which requires long-term rehabilitation and is associated with late-stage complications [47, 48]. The term ‘chronic critical illness’ has been coined to refer to this rapidly increasing proportion of intensive care unit (ICU) survivors that continue to have long term sequelae of their acute illness [49]. Research relating to the changing dynamic of sepsis-related illness is further complicated by a paucity of reliable, long-term outcomes data. This dearth of long-term data disproportionately affects diseases such as sepsis, for which (1) mortality rates are high, (2) hospital coding/administrative data is inconsistent (sepsis definitions having changed twice since their initial iteration in 1992 [20]), and (3) follow-up data is difficult to obtain (patients are difficult to contact due to prolonged, post-hospitalization stays in rehabilitation facilities) [49-52]. Nonetheless, this data is of critical importance to physicians and to their patients, who deserve to understand procedural risks and benefits prior to making truly informed decisions about their healthcare.

It has been reported that muscle wasting comorbidities, rather than sarcopenia itself, predict short-term hospital outcomes following sepsis [17]. In contrast, a recent, larger study examined the impact of sarcopenia on short-and long-term outcomes in patients with septic shock and observed that sarcopenia was associated with increased 28-day (HR 2.1) and long-term (HR 1.7) mortality [53]. Both studies were retrospective in nature, although the latter study included patients with shock, who inherently experience higher mortality rates as compared with patients having sepsis alone [54]. Furthermore, the study by Oh *et al* did not evaluate the impact of sarcopenia on post-hospitalization discharge status, which may serve as a surrogate measure for quality of life in survivors of critical illness. Our study is the first, to our knowledge, to report the effect of sarcopenia on long-term mortality and rehabilitation needs, in critically ill patients with sepsis.

The juxtaposed findings of increased age-adjusted, one-year mortality with unchanged post-hospitalization discharge disposition, in critically ill patients with sepsis and pre-existing sarcopenia is notable. It is plausible that elderly patients with significant medical comorbidities but without pre-existing muscle dysfunction are more likely to require the intensive support provided by skilled nursing services and rehabilitation facilities. Elderly patients are also more likely to request hospice care in the face of a prolonged and uncertain disease course. Clinician ‘intuition,’ however, may also be a confounding variable in the decision to pursue post-hospitalization nursing care in these patients. Clinicians often make assumptions about the frailty of older adults, independent of actual needs [55, 56].

The binary nature of one-year mortality, combined with pre-defined imaging criteria for sarcopenia, may provide more objective insight into the long-term effects of muscle dysfunction in this patient population. Further prospective research is needed to discern the influences of sarcopenia and age on post-hospitalization care.

While “sarcopenia” was a term coined by Rosenberg to indicate loss of muscle mass [57], “frailty” is a multidimensional, geriatric syndrome characterized by reduced homeostatic reserves and increased risk of negative health-related events [58]. Unlike frailty assessments, which require an interview of patients or their families to assess slowness, weakness, weight loss and exhaustion [59, 60], image-based assessments of sarcopenia do not require reliable historic medical information. CT imaging-based risk assessment tools are therefore particularly useful in sepsis, since these patients often undergo imaging as part of their medical work-up and the results are rapidly available within the electronic medical record. This is a critical advantage, since patients with sepsis often suffer from confusion and/or encephalopathy, and they are often admitted emergently to the hospital in the absence of family members who could corroborate their medical history.

Strengths of our study include a complete data set of one-year outcomes for all patients, and the validation of sepsis diagnoses by research investigators, according to contemporary Sepsis-3 criteria. Its main limitations are its retrospective nature, its predominantly white population (reflective of the healthcare system’s catchment area), and its single-center design. The inclusion of critically ill patients with sepsis, a population that experiences high mortality rates and frequent discharge to a rehabilitation facility, may also necessarily limit the generalizability (external validity) of our results. However, this population accounts for a large proportion of the healthcare costs and patient morbidity resulting from sepsis, and it is therefore of interest to epidemiologists, physicians, and healthcare administrators alike [5].

Perioperative sarcopenia research has seen a surge over recent years since it has been proposed as a potentially modifiable risk factor for higher costs and increased mortality/complication rates in post-surgical patients [61]. The reluctance of surgeons to operate on severely debilitated patients combined with ubiquitous sepsis-alert systems in the postoperative period have significantly reduced the incidence of surgical sepsis over the past 15 years [45]. Nevertheless, sarcopenia remains a significant comorbidity in the 30-50% of hospitalized, non-surgical patients who die from sepsis [62, 63]. Future studies may uncover differing effects of sarcopenia in surgical and non-surgical patients with sepsis. A better understanding of the pathophysiology of muscle loss and dysfunction may also shed light on the reversibility of sarcopenia by pre-habilitation and other measures such as early mobilization and nutritional optimization, in patients who are at high risk of experiencing poor clinical outcomes.

## Data Availability

All data produced in the present work are contained in the manuscript

## Acknowledgements

We would like to acknowledge Dr. Vernon Chinchilli for his mentorship and guidance with data analysis.

